# Outcomes Following Pulmonary Vein Isolation in ESRD Patients: USRDS-Based Outcome Data

**DOI:** 10.1101/2025.05.19.25327962

**Authors:** Het Patel, Ashraf Alzahrani, Lillie Lamont, Ayeesha Kattubadi, David A Hamon, Edward M Powers, Sergio Conti, Peter D Farjo, Paari Dominic

**Author notes:** **<u>Principle Investigator:</u>** Paari Dominic, MBBS, MPH, 200 Hawkins Drive, Iowa City, IA 52242. **<u>Ethical Approval:</u>** This study was approved by The University of Iowa Institutional Review Board in accordance with ethical guidelines and regulations. The authors ensured compliance with all relevant data protection and privacy regulations while conducting the research. **<u>Disclosures:</u>** The data reported here have been supplied by the United States Renal Data System (USRDS). The interpretation and reporting of these data are the responsibility of the author(s) and in no way should be seen as an official policy or interpretation of the U.S. Government. **<u>Clinical Question and Findings:</u>** What are the outcomes of PVI ablation in patients with ESRD and atrial fibrillation? PVI ablation in patients with ESRD and a diagnosis of AF was associated with lower overall mortality but is associated with a higher rate of hospitalization.

## Abstract

**Background:** In End-Stage Renal Disease (ESRD) patients, the coexistence of AF significantly elevates the risks of stroke (4.8 - 5.6 per 100 person-years) and mortality (26.9 per 100 person-years). Current AF literature lacks insights into pulmonary vein isolation (PVI) outcomes in this population. We aimed to assess the outcomes of PVI in ESRD patients with AF.

**Methods and Materials:** We included all adult patients with a pre-existing diagnosis of ESRD and at least one prior admission for AF from the United States Renal Data System (USRDS) registry and stratified them according to Pulmonary Vein Isolation (PVI). Those with prior transplantation were excluded. Kaplan-Meier curves were used for time-to-evet analysis. Logistic regression analysis was used to assess hazard ratios. Propensity score analysis was used to evaluate the treatment effect after adjusting for covariates.

**Results:** The study included 103,982 ESRD patients with AF, of whom 4,314 (4.15%) received ablation, and 99,668 (95.85%) were managed medically. After matching, there was a reduction in mortality (90.2% vs 93.4%, p<0.001) among the ablation group compared to no ablation at 5 years. Increased age (60-69 years and ≥70 years) was associated with 3-fold and 6-fold higher odds of mortality, respectively. A history of congestive heart failure increased the odds of mortality by 36%, while prior stroke, coronary disease, and cancer did not.

**Conclusion:** In ESRD patients with AF, despite a high 5-year mortality, those who undergo PVI exhibit a lower mortality rate compared to those managed without PVI.

## Introduction

Atrial Fibrillation (AF) exhibits a global prevalence ranging from 11% to 27%.^1–3^ Analyzing data from the United States Renal Data System (USRDS) spanning 1989 to 2006, we observed a threefold increase in the prevalence of AF among End-Stage Renal Disease (ESRD) patients, rising from 3.5% to 10.6%.^1^ The coexistence of ESRD and AF significantly amplifies the risks of stroke and mortality, with respective incidences of 4.8 - 5.6 per 100 person-years and 26.9 per 100 person-years.^4–6^ These rates starkly contrast with ESRD patients without AF, who experience lower incidences of 1.9 and 13.4 per 100 patient-years for stroke and mortality rates respectively.^6^

Registry data from the ORBIT-AF study suggests that all-cause mortality remains unaffected by anti-arrhythmic drugs in patients with AF.^7^ In contrast, catheter ablation has emerged as a promising alternative, demonstrating superior rhythm control compared to anti-arrhythmic drugs.^8^ Notably, catheter ablation may be associated with improved outcomes, particularly in reducing deaths secondary to heart failure and maintaining sinus rhythm.^9,10^ Moreover, successful catheter ablation has demonstrated positive effects on left ventricular contractility, increased cardiac output, and improved renal function in chronic kidney disease (CKD) patients with cardio-renal syndrome.^11–13^ However, literature on the effects of catheter ablation in ESRD remains limited as this population has been notably excluded from clinical trials. ^6,14^ A meta-analysis by Zimmerman reported a reluctance among electrophysiologists to pursue catheter ablation in ESRD patients with AF.^6^

Given the existing gap in current AF guidelines regarding outcomes specific to catheter ablation in patients with ESRD and AF, we sought to address this knowledge deficit. Our study aims to evaluate the outcomes of ESRD patients with AF after undergoing catheter ablation compared to those managed without ablation. Additionally, our investigation extends to the exploration of risk factors associated with mortality in ESRD patients with AF.

## Methods and materials

Utilizing hospitalization data within the USRDS registry, adult patients with a pre-existing diagnosis of ESRD who were admitted with AF were included. Those who had undergone kidney transplant previously and those with missing information were excluded. Subsequently, the cohort was stratified into two groups: those who received catheter ablation and those who were managed without ablation.

### Data abstraction

Utilizing the USRDS registry, we conducted a data integration process. The Medevid file and other pertinent files containing information on comorbidities and mortality were merged, employing the unique USRDS identification number as the linking variable. The date of AF diagnosis was extracted from the claim “from” and “through” variables in the hospitalization file. Time to ablation was calculated by measuring the duration from the date of AF diagnosis to the specific date of ablation, as indicated by the claim “from” variable. Comprehensive data on comorbidities, mortality, and date of death were identified using the Medevid file. Time to death was measured from the date of AF diagnosis to the date of death for the group without ablation and from the date of ablation to the date of death for patients who underwent ablation.

### Outcomes

The primary outcome was mortality. The secondary outcomes were mortality due to cardiac causes, hospitalization, and hospitalization for cardiac causes. The follow-up duration was up to 5 years from the index admission for AF.

### Statistics

Descriptive statistics were used to compare baseline characteristics. Propensity score matching was performed to achieve a 1:1 matching ratio, with means and proportions compared between groups post-matching. Logistic regression and odds ratios were assessed before and after propensity score matching. Treatment effects were analyzed following propensity matching. Time-to-event analysis was conducted using Kaplan-Meier curves and log-rank tests. A significance level of p<0.05 was adopted for this study. All statistical analyses were performed using STATA software version 17.1 SE.

### Ethical considerations

This study was approved by The University of Iowa Institutional Review Board in accordance with ethical guidelines and regulations.

## Results

A total of 103,982 patients were included, with 4,314 (4.15%) receiving ablation and the remaining 99,668 (95.85%) managed medically. The median age at diagnosis was notably higher in the no-ablation group compared to the ablation group (75 years vs. 68 years, p<0.001), and a higher proportion of female patients were observed in the no-ablation group (40.8% vs. 36.4%, p<0.001). The ablation group exhibited a higher representation of African American race and cancer. In contrast, the no-ablation group had a higher proportion of individuals identifying as white or Asian, as well as a higher prevalence of congestive heart failure (CHF) and transient ischemic event (TIA)/cerebrovascular accident (CVA) (Table 1).

**Table 1:** Baseline Characteristics Before and After Propensity Score Matching.

|  | No Ablation<br>(n=99,968) | Ablation<br>(n=4,314) | p-value | No Ablation<br>(n=4,314) | Ablation<br>(n=4,312) | p-value |
| --- | --- | --- | --- | --- | --- | --- |
| Age (mean, SD)<br>years | 73.8 (SD: 10.4) | 67.4 (SD: 11.5) | <0.001 | 68.5 (SD:<br>12.4) | 67.4 (SD: 11.5) | 0.98 |
| Gender (Female) | 40,671<br>(40.8%) | 1572 (36.4%) | <0.001 | 1571 (36.4%) | 1570 (36.4%) | 0.98 |
| Race: |  |  |  |  |  |  |
| White | 77,071<br>(77.3%) | 2737 (63.4%) | <0.001 | 2735 (63.4%) | 2737 (63.5%) | 0.94 |
| African American<br>(AA) | 18,187<br>(18.2%) | 1441 (33.4%) | <0.001 | 1364 (31.6%) | 1439 (33.4%) | 0.08 |
| Asian | 2985 (2.9%) | 80 (1.8%) | <0.001 | 124 (2.8%) | 80 (1.8%) | 0.002 |
| Other | 1428 (1.4%) | 56 (1.3%) | 0.46 | 91 (2.1%) | 56 (1.3%) | 0.004 |
| **Comorbidities** |  |  |  |  |  |  |
| CHF | 44,950<br>(45.1%) | 1722 (39.9%) | <0.001 | 1723 (39.9%) | 1722 (39.9%) | 1.00 |
| COPD | 14,015<br>(14.0%) | 528 (12.2%) | 0.001 | 292 (6.2%) | 528 (12.2%) | <0.001 |
| Cancer | 8693 (8.7%) | 317 (7.3%) | 0.002 | 316 (7.3%) | 317 (7.3%) | 0.96 |
| TIA/CVA | 10,555<br>(10.6%) | 355 (8.2%) | <0.001 | 357 (8.3%) | 355 (8.2%) | 1.00 |
| CAD | 24,445<br>(24.5%) | 923 (21.4%) | <0.001 | 924 (21.4%) | 923 (21.4%) | 0.98 |
| Total<br>Hospitalizations | 5 (3-8) | 9 (6-14) | <0.001 | 5 (3-8) | 10 (6-15) | <0.001 |
| Hospitalizations due<br>to cardiac issues | 0 (0-1) | 2 (1-4) | <0.001 | 0(0-1) | 2 (1-4) | <0.001 |
| Stroke after<br>diagnosis | 5039 (5.1%) | 280 (6.5%) | <0.001 | 245 (5.7%) | 280 (6.5%) | 0.11 |
| **Outcomes** |  |  |  |  |  |  |
| Overall mortality | 91,600<br>(91.9%) | 3,892 (90.2%) | <0.001 | 4,031 (93.4%) | 3,892 (90.2%) | <0.001 |
| Mortality due to<br>cardiac issues | 32,482<br>(32.6%) | 1589 (36.8%) | <0.001 | 1487 (34.5%) | 1589 (36.8%) | 0.02 |
| Time to Death<br>(days) | 515 (181–<br>1094) | 1029(514-1693) | <0.001 | 820 (± 782) | 1166 (±788) | <0.001 |

The overall mortality was 91.9% in the no ablation group compared to 90.2% in the ablation group (p<0.001) at 5 years (Figure 1(a)). Patients lived longer in the ablation group compared to no ablation group (1019 days vs 515 days, p<0.001). The number of hospitalizations was higher in the ablation group compared to no ablation group (5 vs 9, p<0.001) at 5 years. Similarly, hospitalizations from cardiac causes were significantly higher in the ablation group compared to no ablation group (0 vs 2, p<0.001) at 5 years. Before propensity matching, there was a statistically significant difference in stroke in the ablation group compared to no ablation (6.5% vs 5.1%, p<0.001). However, this observed increase in stroke rate in the ablation group was no longer present after propensity matching.

**Figure 1(a):**
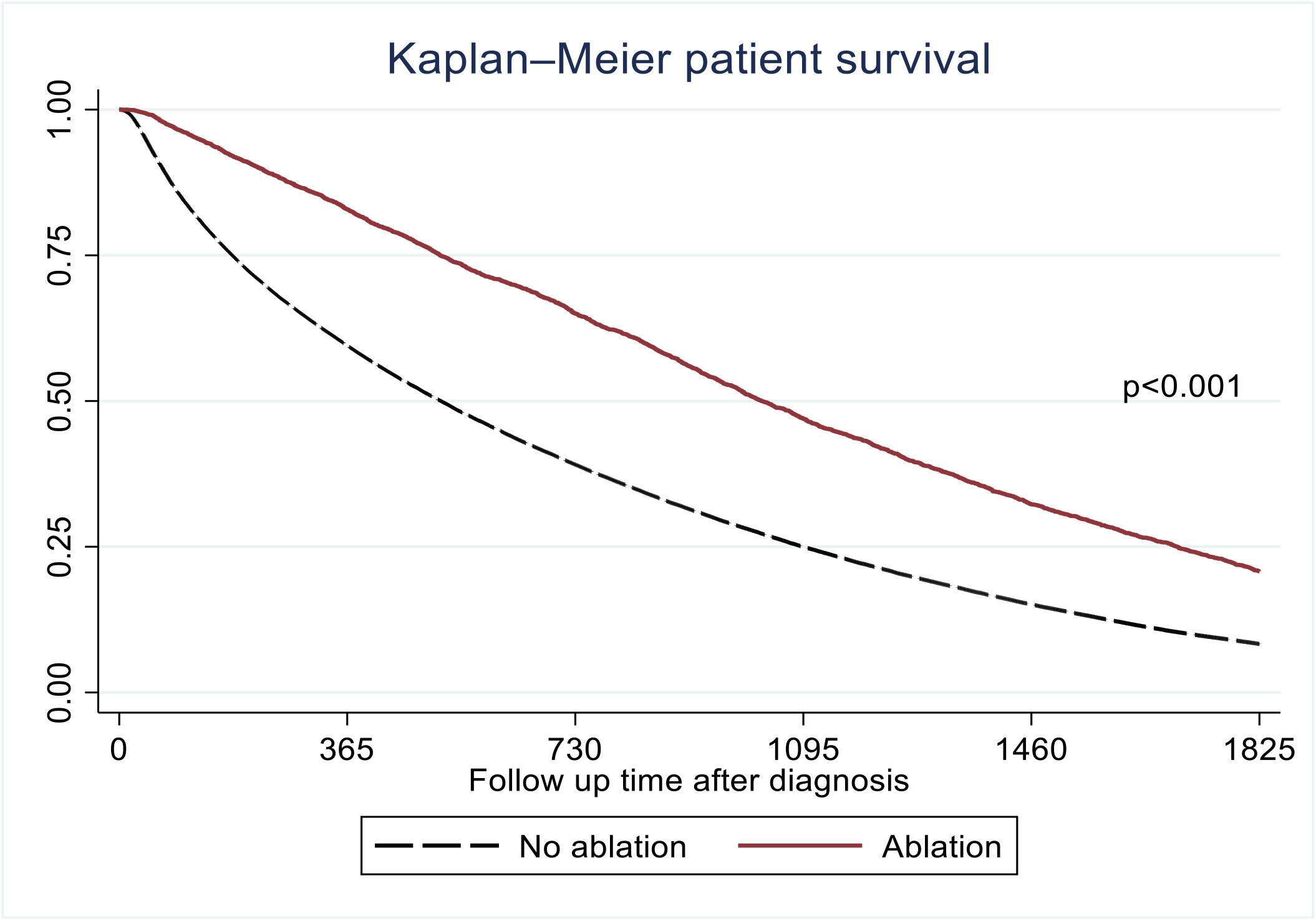
Kaplan Meier Survival Curve Comparing PVI vs no PVI in AF Patients with ESRD Before Propensity Score matching.

**Figure 1(b):**
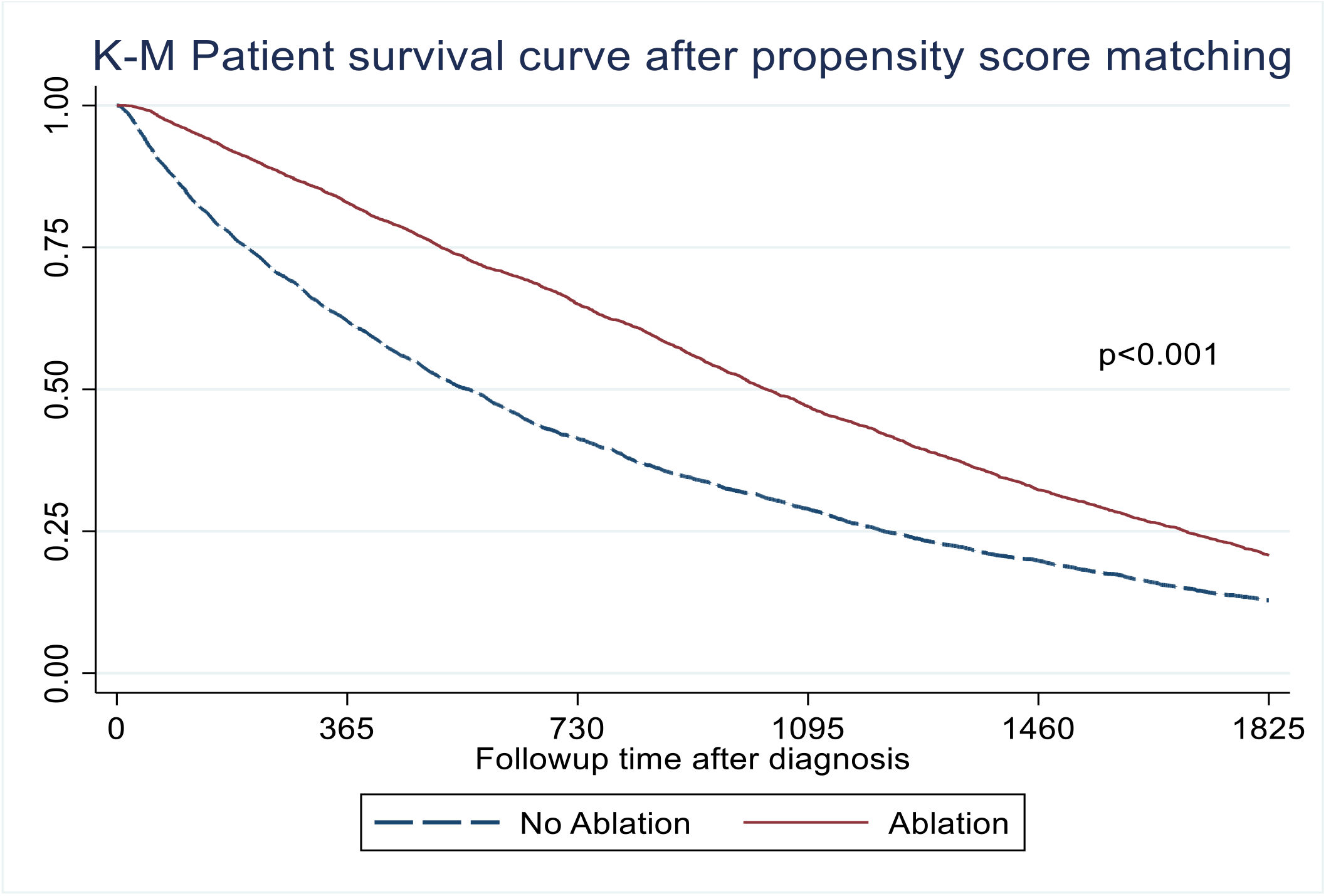
Kaplan Meier Survival Curve Comparing PVI vs no PVI in AF Patients with ESRD After Propensity Score matching.

Propensity score matching in a 1:1 fashion allowed for more comparable baseline characteristics between the two groups. Specifically, we achieved an adequate match in age, gender, white race, and all comorbid conditions except COPD. A detailed comparison of the matched groups is provided in Table 1. Following propensity matching, there remained a significant difference in mortality rate between the groups (93.4% in the no ablation group vs 90.2% in the ablation group, p<0.001) (Figure 1(b)), translating into a 3.2% absolute reduction in mortality in the ablation group. In addition, time to death was delayed by 346 days among the patients who received ablation compared to no ablation. The ablation group had more hospitalizations (10 vs 5, p<0.001) and hospitalizations due to cardiac cause (2 vs 0, p<0.001). As discussed, there was no difference in the stroke rate between both groups after propensity matching (5.7% vs 6.5%, p=0.11).

Logistic regression was used to analyze factors contributing to increased mortality among patients who suffered AF irrespective of ablation before and after propensity matching. Notably, the odds ratio before and after matching differed significantly. After matching, advanced age was associated with a 3-fold and 6-fold increase in mortality for those aged 60-69 years and Age ≥70 years, respectively. Furthermore, a history of CHF increased the odds of mortality by 36%. However, prior CVA, coronary artery disease (CAD), and cancer were not associated with increased odds of mortality after propensity matching (Table 2, Figure 2).

**Table 2:** Logistic Regression with Odds Ratio Before and After Propensity Score Matching.

| Before Propensity score matching |  |  | After Propensity score match |  |
| --- | --- | --- | --- | --- |
|  | Odds ratio (95% CI) | p-value | Odds ratio (95% CI) | p-value |
| Age 18-39 years | Reference | --- | Reference | ---- |
| Age 40-59 years | 1.88 (1.53 – 2.31) | <0.001 | 2.21 (1.45 – 3.38) | <0.001 |
| 60-69 | 2.85 (2.33 – 3.50) | <0.001 | 3.43 (2.23 – 5.26) | <0.001 |
| >=70 | 6.31 (5.15 – 7.72) | <0.001 | 6.77 (4.37 – 10.5) | <0.001 |
| Gender (Female) | 1.02 (0.97 – 1.07) | 0.34 | 1.16 (0.98 – 1.37) | 0.09 |
| White | 1.61 (1.53 – 1.69) | <0.001 | 1.55 (1.31 – 1.82) | <0.001 |
| CHF | 1.60 (1.52 – 1.68) | <0.001 | 1.36 (1.14 – 1.62) | <0.001 |
| CAD | 1.22 (1.14 – 1.29) | <0.001 | 1.22 (0.96 – 1.53) | 0.096 |
| Cancer | 1.21 (1.10 – 1.33) | <0.001 | 1.49 (0.99 – 2.25) | 0.052 |
| CVA | 1.15 (1.06 – 1.25) | <0.001 | 0.97 (0.72 – 1.31) | 0.85 |

**Figure 2:**
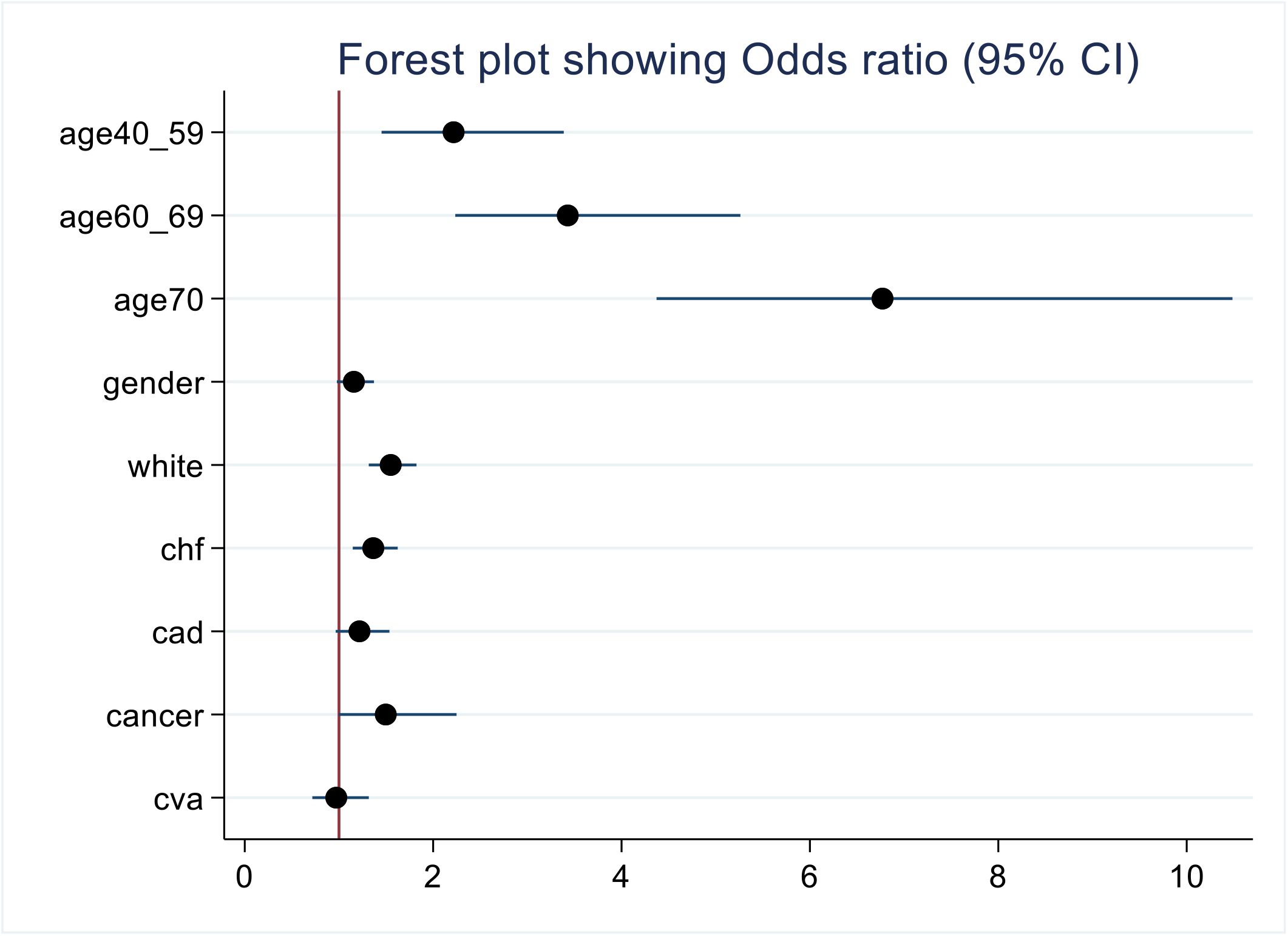
Forest Plot of Odds Ratios and 95 % CIs for Factors Associated with Mortality in AF Patients with ESRD who Undergo PVI.

The overall 5-year survival rate was poor in ESRD patients suffering from AF (<25%). However, the ablation group showed a statistically significant improvement in survival following ablation (p<0.001), Figure 1. This holds true after propensity matching. (Table 2).

## Discussion

Comorbid AF in patients with ESRD is associated with an increase in morbidity and mortality. Despite this, patients with ESRD remain underrepresented in AF ablation trials, and there is often a reluctance amongst providers to pursue catheter ablation therapy in this group. This retrospective analysis of USRDS data provides real-world insights into the outcomes of PVI in patients with ESRD.

Most notably, our analysis demonstrated a reduction in overall mortality with catheter ablation that persisted after propensity score matching (93.4% in the no ablation group vs. 90.2% in the ablation group, p<0.001), translating into a 3.2% absolute reduction in mortality in the ablation group. In addition, on average, patients in the ablation group lived one year longer (1166 days vs 820, p<0.001). In the general population, clinical trials have shown mixed results with regards to outcomes after an AF ablation^10, 15^. However, subsequent studies have shown that select high-risk patients, specifically those with established CHF, may derive a mortality benefit from catheter ablation.^9,16^ This may hold true for patients with ESRD, a group known to have highly prevalent co-existent cardiovascular disease, including heart failure.^17^ Indeed, our logistic regression analysis demonstrated that CHF was associated with a 36% increase in mortality in those who underwent catheter ablation. It may be that the apparent benefit derived from catheter ablation in this cohort can be attributed to the improvement in cardiac function and left ventricular ejection fraction associated with the maintenance of normal sinus rhythm^18,19^. Furthermore, restoring normal sinus rhythm has been shown to improve renal function in patients with CHF and CKD suffering from cardiorenal syndrome.^11^

Impaired renal function has been shown to predict early recurrence of AF. ^20^ In a study by Yanagisawa et al., the overall recurrence rate was 18% after catheter ablation during nine months of follow-up, with subgroup analysis showing a 46% recurrence rate in patients with stage 3 CKD. It may come as no surprise that a metanalysis by Zimmerman et al. reported that 37.6% of cardiologists in major university-based hospitals in the USA would not consider catheter ablation for patients if the creatinine clearance were <30 mL/min in non-dialysis patients and 11.3% would not consider AF ablation in patients on dialysis.^6^

In this study, the number of overall hospitalizations (10 vs 5, p<0.001) and hospitalizations due to cardiac cause (2 vs 0, p<0.001) was higher in the ablation group. This may be attributed to various factors such as increased survival time, allowing for more hospitalizations for atrial fibrillation recurrence, heart failure decompensation, or dialysis-related complications.

Alternatively, patients who undergo ablation may be more likely to be admitted for additional rhythm control strategies in cases of suboptimal response to ablation. Another explanation for the increased hospitalizations may be complications associated with the catheter ablation procedure itself, such as vascular access issues, bleeding, pericarditis, or pericardial effusion. The increase in hospitalizations among the ablation group suggests the need for further investigation into the complications associated with this intervention.

## Strengths & Limitations

This study provides insights into the impact of PVI on mortality in ESRD patients, a population often excluded from related trials. The USRDS database provides real-world data, enhancing the generalizability of our findings and the large sample size allows for a comprehensive analysis of the impact of PVI on outcomes.

While our study provides valuable insights, it is important to acknowledge several limitations inherent in using USRDS data. In general, as with many retrospective studies, this study has inherent limitations, including the missing or incomplete data. Patients who undergo ablation in general may be healthier compared to the patients who did not; we attempted to control this confounding through propensity matching and achieved reasonably comparable groups.

Furthermore, the absence of pertinent laboratory values at the time of the procedure and the lack of detailed information on echocardiography findings during any point in the patient’s history are notable limitations.

## Conclusion

In ESRD patients with AF, the 5-year overall mortality remains high. Still, those who underwent PVI exhibited a lower mortality rate compared to those managed without PVI but with a higher rate of hospitalization. This underscores the potential benefits of catheter ablation in improving outcomes for this high-risk patient population.

## Data Availability

This study is based on national renal registry data base called USRDS.

https://www.niddk.nih.gov/about-niddk/strategic-plans-reports/usrds

